# THE IMPLEMENTATION AND OUTCOME OF DIABETES PREVENTION PROGRAM LIFESTYLE INTERVENTION IN HIGH RISK OR PREDIABETES: A REVIEW OF THE LITERATURE

**DOI:** 10.1101/2023.10.12.23296982

**Authors:** Oliva Suyen Ningsih, Moses Glorino Rumambo Pandin, Ferry Efendy

## Abstract

**Background:** Effective lifestyle intervention can lower Type 2 diabetes mellitus in high-risk or prediabetic people through diabetes mellitus prevention programmes. The aim of this review of the research is to provide an explanation of how diabetes prevention programmes (DPP) are implemented in high-risk people or those with prediabetes using a variety of DPP methods and results.

**Method:** The method used is a literature review by searching for relevant peer-reviewed articles published in English in three databases: Scopus, PubMed, and CINAHL. Studies assessing the use of diabetes mellitus preventive programmes in high-risk people or people with prediabetes were the inclusion criteria for this review. Individuals with prediabetes or high risk were among the subjects. The main result is the outcome of the DPP. Literature search is limited to publications in 2018–2023 and open access.

**Result:** 18 articles in all were reviewed.The main focus of DPP-based lifestyle interventions is dietary and physical activity. Health education and behaviour change strategies are also included in lifestyle interventions. The implementation of nutrition or dietary interventions can be provided face-to-face on a group-based basis for an average of 6–12 months and online through intensive interventions with weekly online modules through web-based platforms, text messaging, and digital therapy. The implementation of diet and physical activity interventions needs support from families or groups to be able to make healthy behaviour changes, one of which is peer support. The short-term effects of DPP are decreased waist circumference, plasma glucose levels, increased knowledge, diet, and physical activity. The long-term consequences of DPP include weight loss with a lowered risk of developing diabetes, modifications to lipid profiles, enhanced pancreatic cell function, increased insulin sensitivity, decreased body fat mass, and HbA1C.

**Conclusion:** Diabetes mellitus prevention programme through effective lifestyle interventions to slow the onset of type 2 diabetes mellitus in prediabetic individuals

## BACKGROUND

Diabetes mellitus is a non-communicable disease that can cause death (WHO, 2023). The International Diabetes Federation (IDF) estimates that 573 million adult people (20-79 years old) worldwide already have diabetes, and that number will rise to 643 million by 2030 and 783 million by 2045 (IDF, 2021). Microvascular consequences of type 2 diabetes Mellitus might include retinopathy, nephropathy, and neuropathy (Thipsawat, 2023). Prediabetes is one factor that can accelerate the onset of diabetes mellitus (Venkataramani et al., 2019). According to Rooney et al. (2023), prediabetes is characterised by impaired glucose tolerance (IGT) (2-h glucose [140–199 mg/dL]) and impaired glucose tolerance (IFG) (fasting glucose [110–125 mg/dL]) or increased blood glucose levels below the threshold for a diagnosis of diabetes. Approximately 96 million persons aged 18 and over were expected to have prediabetes in 2019 by the Centres for Disease Control and Prevention (CDC, 2022). According to Rooney et al. (2023), those with IGT and IFG have a high chance of getting diabetes within 5 years (up to 50%). Diabetes prevention through lifestyle interventions needs to be carried out in Prediabetic individuals to slow the onset of diabetes mellitus (Jiang et al., 2018).

According to several studies, lifestyle interventions can prevent 40–70% of prediabetic people from developing Type 2 diabetes mellitus (Kramer et al., 2018; Sampson et al., 2020; Thipsawat, 2023; Vargas et al., 2023). The main focus of DPP through lifestyle interventions is diet and physical activity (Ford et al., 2019; Kriska et al., 2021). Food composition is an integral part of diabetes mellitus management by considering three components, namely encouraging weight loss or maintenance, improving glycemic control, and preventing complications (Hall, Rosemary; Krebs & Parry Strong, 2016). Low-carbohydrate diets are effective in reducing weight and improving glycemic control (Brouns, 2018). Diabetes mellitus can be prevented by engaging in moderate-intensity physical activity for 150 minutes five days a week or longer, such as brisk walking, two to three sessions of muscle-strengthening exercises, and lowering total and saturated fat intake (Kramer et al., 2018; Sampson et al., 2020).

All participants in the DPP lifestyle intervention receive the same intervention to help them lose weight and get more exercise, but they are also free to choose how they want to go about accomplishing it (American Diabetes Association, 2021). Previous literature studies only provide a general description of the recommended components of diet and physical activity (MacPherson et al., 2020; Muilwijk et al., 2018), but the literature review describes the implementation of DPP through lifestyle interventions with various methods supported by limited evidence of its effectiveness. The aim of this literature review is to describe how the DPP lifestyle intervention is implemented the outcome in high-risk people or people with prediabetes.

## METHOD

The method used is literature-review by searching for relevant peer-reviewed articles published in English in three databases: Scopus, PubMed, and CINAHL. Studies assessing the application of diabetes mellitus preventive interventions in people at high risk of developing diabetes mellitus or prediabetes were the inclusion criteria for this evaluation. The types of research are observation, experiment, randomized controlled trial, non-randomized controlled trial. Individuals with prediabetes or diabetes mellitus were among the subjects. The main result is the outcome of the DPP. Open access and 2018-2023 publications alone are included in the literature search.

## RESULT

The author found 34,319 articles from three databases, including Scopus: 701 articles; Pubmed: 16,866 articles; CINAHL: 16,752 articles, after searching with specific keywords combined with Boolean “AND” and “OR” operators. The keywords of the PICO framework include Population (individuals at high risk of Diabetes mellitus or Prediabetes), Intervention (Diabetes Prevention), Comparison (usual care), and Results (Risk of diabetes mellitus or clinical condition). 18 papers were assessed after being duplicate-screened by Rayyan and manually by the research team while taking the inclusion criteria into consideration.

**Figure 1.**
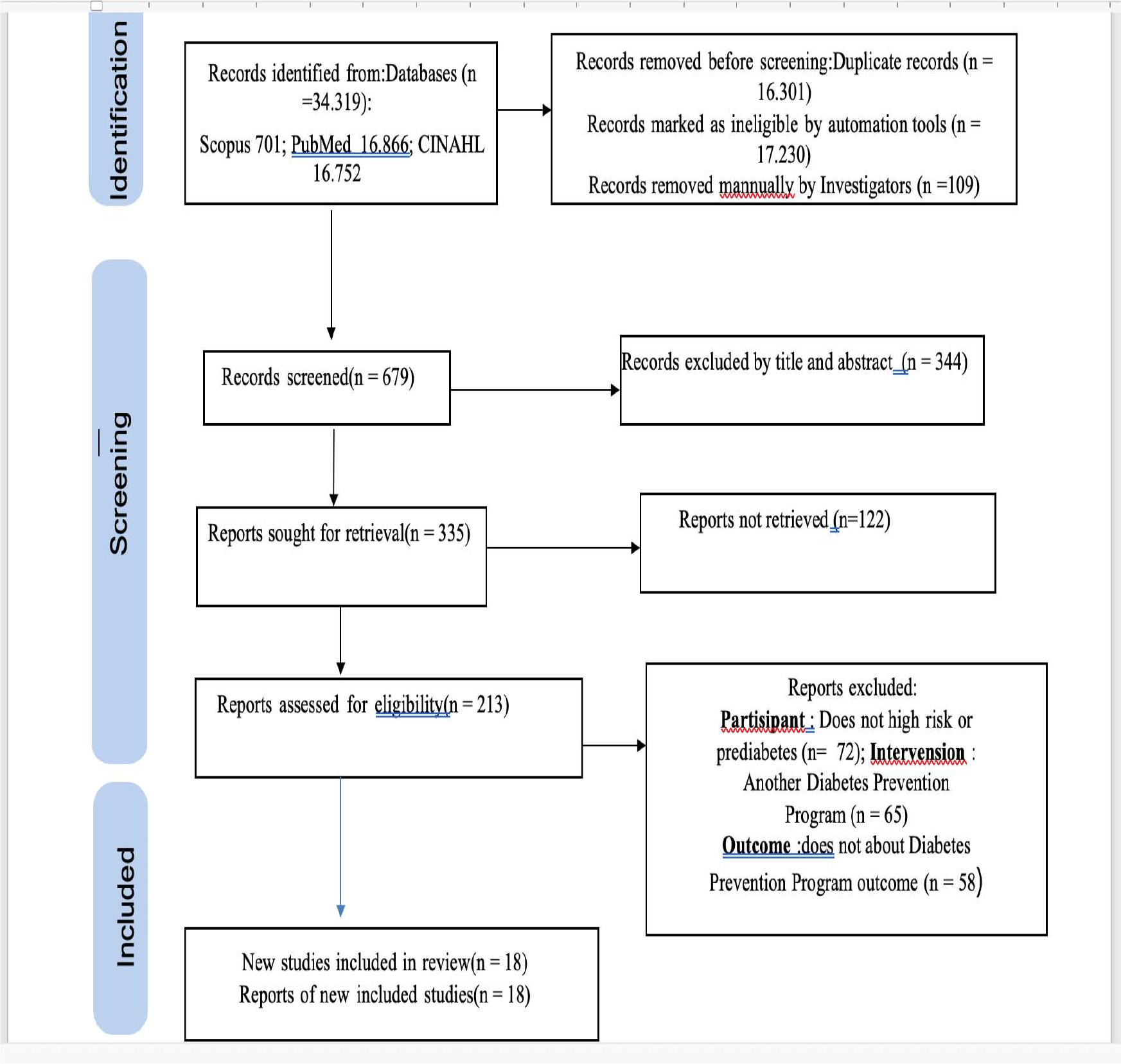
Flowchart for PRISMA

**Table 1.** Characteristics of reviewed studies.

| NO | Title, author,year | Method | Result |
| --- | --- | --- | --- |
| 1 | “Older Adults and Diabetes Prevention Programs in the Veterans Health Administration “ (Lee, Pearl G; Damschroder, Laura J; Holleman, Robert; Moin, Tannaz; Richardson, 2018) | **Desain** : nonrandomized comparative effectiveness trials<br>**Subjects**: respondents aged ≥ 65 years = 120, < 65 years = 258 respondents.<br>**Variable** :<br>**Independent** :Diabetes prevention program<br>**Dependent** : weight loss<br>**Instruments**: questionnaire<br>**Analysis** : ANOVA | The online diabetes mellitus prevention intervention group participated more than the African veterans department diabetes mellitus prevention intervention group (P < 0.05). Both groups experienced a 5% weight loss after the intervention. |
| 2 | “Design and rationale of behavioral nudges for diabetes prevention (BEGIN): A pragmatic, cluster randomized trial of text messaging and a decision aid intervention for primary care patients with prediabetes “(Vargas et al., 2023)” | **Design**: cluster randomized trial<br>**Subjects**: 656 Prediabetic Individuals<br>**Variable**:<br>**Independent** : behavioral nudges for diabetes prevention (BEGIN):<br>**Dependent**: weight loss , adoption of diabetes preventive care, hemoglobin A1c<br>**Instrument**: electronic Health Record, questionnaire | Weight loss in 12 months, implementation of evidence-based treatment to prevent diabetes and improvement in hemoglobin A1c. |
|  |  | **Analysis** : Descriptive statistics, linear regression |  |
| 3 | “Combining supervised run interval training or moderate□intensity continuous training with the diabetes prevention program on clinical outcomes” (Gilbertson et al., 2019)” | **Desain** : randomized controlled trial<br>**Subjects:** 37 adults, aged 18-71 years<br>**Variable:**<br>**Independent** : Diabetes prevention program (DPP) combined with run sprint interval training (INT) or moderate-intensity continuous training/MICT)<br>**Dependent:**. glycemic control, body composition, fitness, exercise adherence, and enjoyment of exercise<br>**Instrument** : questionnaire<br>**Analysis** : ANOVA, Wilcoxon, Mann–Whitney | The moderate-intensity continuous training (MICT) group felt more enjoyment when exercising. Both groups experienced a decrease in body weight, BMI, body fat mass, fasting glucose and HbA1c. The MICT experienced greater reductions in weight, BMI, and body fat than the INT group. |
| 4. | “Evaluation of a Diabetes Prevention Program Lifestyle Intervention in Older Adults “(Kramer et al., 2018)” | **Desain** : randomized controlled study<br>**Subjects** : 134 Adult individuals with prediabetes or metabolic syndrome, BMI ≥24 kg/m2<br>**Variable:**<br>**Independent** : Diabetes Prevention Program (DPP) lifestyle program<br>**Dependent:**. Weight loss, A1C, fasting blood sugar, waist circumference, physical activity, total cholesterol, HDL, LDL, Triglycerides, systolic blood pressure.<br>**Instrument** : questionnaire<br>**Analysis** : 2-sampel t test , Paired t test dan wilxon | At 6 months, the intervention group showed greater average weight loss than the control group as well as much greater improvements in physical activity, A1C, fasting blood sugar and waist circumference. In *the pre-post analysis*, both groups of randomized showed the same success that was maintained at 18 months. |
| 5 | “Clinic- and Community-Based National Diabetes Prevention Programs in Los Angeles “ (Defosset et al., 2021)” | **Desain** : clinic- versus community-based setting.<br>**Subjek** : 265 respondents<br>**Variable:**<br>**Independen** : *Diabetes Prevention Program*<br>**Dependent:**.presence, physical activity, weight<br>**Instrument** : Questionnaire<br>**Analysis** : Negative binomial regression models tested | Most respondents were Hispanic/Latino (81.51%) and female (90.19%). Programs implemented by CBOs (*Clinics and community-based organizations*) involved other racial/ethnic groups including more black participants than clinics (20.56% vs 0%); more men were involved (15.29% vs 7.22%). Some participants had met weight loss goals under the National *National Diabetes Prevention Program* (DPP) (clinics: 15.29%, CBO: 20.00%). |
| 6 | “Effect of a family and interdisciplinary intervention to prevent T2D: randomized clinical trial” (Vargas-ortiz et al., 2020)” | **Design:** Randomized Clinical Trial<br>**Subject;** 122 respondents with prediabetes, and 101 family members<br>**Variable:**<br>**Independent** : a family and interdisciplinary intervention<br>**Dependent:** glucose, insulin resistance, pancreatic β cell function, lipid profile and anthropometric<br>**Instrument** : Questionnaire<br>**Analysis** : independent t test and ANOVA | Family members who received the *interdisciplinary family intervention* experienced changes in the glucose curve (from $18.597 \pm 2611$ to $17.237 \pm 2792$, $p = 0.004$) and insulin sensitivity/*matsuda index* (from $3.5 \pm 2.3$ to $4.7 \pm 3.5$, $p = 0.05$) after 12 months. The individual intervention group had an increase in the Disposition Index/function of Pankeas cells (from $1.5 \pm 0.4$ to $1.9 \pm 0.73$, $p < 0.0001$) after 12 months. There was improvement in body weight and lipid profile at 6 months and no improvement in the individual intervention group at 12 months while in the *interdisciplinary family* group there was still improvement. Adherence of up to 12 months did not differ between *the interdisciplinary family* group and the individual intervention (FI 56% Vs II 60%). |
| 7 | “Lifestyle Intervention With or Without Lay Volunteers to Prevent Type 2 Diabetes in People With Impaired Fasting Glucose and/or Nondiabetic Hyperglycemia A Randomized Clinical Trial” (Sampson et al., 2020)” | **Design:** randomized clinical trial<br>**Subject :** 1028 respondents<br>**Variable:**<br>**Independent :** Group-based lifestyle interventions<br>**Dependent :** HbA1c, fasting blood glucose<br>**Instrument : Questionnaire**<br>**Analysis :,** binary logistic regression t test, linear regression | In this study, 1028 participants were randomized (Lifestyle intervention group (INT), 424 [41.2%] [166 women (39.2%)]); Lifestyle intervention group with additional telephone support (INT-DPM), 426 [41.4%] [147 women (34.5%)]; *usual care* (CON), 178 [17.3%] [70 women (%39.3)]) between January 1, 2011, and February 24, 2017. Mean age (SD) 65.3 (10.0) years, average (SD) body mass index 31.2 (5) (calculated as weight in kilograms divided by height in meters squared), and mean (SD) *follow-up* 24.7 (13.4) months. A total of 156 participants progressed to type 2 diabetes, consisting of 39 of 171 receiving *usual care* (22.8%), 55 of 403 receiving INT (13.7%), and 62 of 414 receiving INT-DPM (15.0%). There were no significant differences between the intervention groups in primary outcomes (odds ratio [OR], 1.14; 95% CI, 0.77-1.7; *P* = 0.51), but each intervention group had a significantly lower likelihood of type 2 diabetes (INT: OR, 0.54; 95% CI, 0.34-0.85; *P* = 0.01; INT-DPM: OR, 0.61; 95% CI, 0.39-0.96; *P* = 0.033; combined: OR, 0.57; 95% CI, 0.38-0.87; *P* = 0.01). |
| 8 | “Prevalence and Correlates of Diabetes Prevention Program Referral and Participation”(Venkataramani et al., 2019)” | **Design:** cross-sectional survey<br>**Subject :** 2341 respondents<br>**Variable:**<br>**Independent :** Diabetes prevention program<br>**Dependent:**Referral, Participation, and Interest<br>**Instrument :** Survey<br>**Analysis :** logistic regression analyses. | The study population consisted of 2,341 adults. The majority were female (63%), white (74.6%), non-Hispanic (83.4%), and ≥45-year-old (68.2%). A total of 4.2% reported having been referred to a prevention program for 12 months and only 2.4% reported having participated. In multivariable logistic regression, race correlated with referrals (black and Asian adults were more likely to be referred. To prevention programs) and age were positively correlated with participation. More than 25% of adults who have never been referred or participated report interest in becoming involved in a Diabetes mellitus prevention program. |
| 9 | “The role of Sociodemographic factors on goal achievement in a community-based diabetes prevention program behavioral lifestyle intervention” (Devaraj et al., 2021)” | **Design:** *randomized clinical trial*<br>**Subject :** 240 respondents<br>**Variable:**<br>**Independent :** *Diabetes prevention program*<br>**Dependent:** Physical activity and weight<br>**Instrument :** Questionnaire<br>**Analysis :** *Fisher's Exact* or *Chi-Square tests*. Wilcoxon Signed-Rank tests, Wilcoxon Two-Sample or Kruskal-Wallis tests and multiple regression analysis | The majority of the individuals significantly reduced their weight. Comparatively to non-Hispanic black individuals, non-Hispanic white participants lost more weight (5%) at 6 and 12 months. The people who were retired had the best results with weight loss. At baseline and after a year, men, those with college degrees, and people with higher salaries had a higher likelihood of achieving their exercise goals. |
| 10 | “Dietary changes in a diabetes prevention intervention among people with prediabetes: the Diabetes Community Lifestyle Improvement Program trial” (Ford et al., 2019)” | **Design:** *randomized clinical trial*<br>**Subject :** 485 respondents<br>**Variable:**<br>**Independent :** *Diabetes prevention intervension*<br>**Dependent:** Diet and risk of developing diabetes mellitus<br>**Instrument :** Questionnaire<br>**Analysis :** *Chi-Square tests*, *t test*, *linear regression* | At six months after the intervention, there was an increase in the consumption of fruits and vegetables and a decrease in the overall amount of calories consumed. Consuming fruit and vegetables significantly lessened the impact "(12.2%; *P* = 0.015)" on the intervention group, which was half less likely to have diabetes at one year. |
| 11 | “Effects of a Community-Based Diabetes Prevention Program for Latino Youth with Obesity: A Randomized Controlled Trial “ (Soltero et al., 2018)” | **Design:** *randomized clinical trial*<br>**Subject:** 160 respondents<br>**Variable:**<br>**Independent :** *Community-Based Diabetes Prevention Program*<br>**Dependent:** insulin sensitivity, quality of life, percentage of body fat, waist circumference and BMI<br>**Instrument :** Questionnaire<br>**Analysis :** Mplus 7.0 | Adolescents in the intervention group after 3 months showed significant improvements in insulin sensitivity “(P < 0.05)” and weight-specific quality of life “(P < 0.001)”, and decreased BMI, waist circumference, and body fat compared with controls . Improvements in weight-based quality of life and reductions in BMI and body fat remained significant at 12 months "(P<0.001)", whereas changes in insulin sensitivity did not occur. In the subsample of adolescents with prediabetes, insulin sensitivity "(P=0.01)", weight-based quality of life "(P<0.001)", and BMI "(P<0.001)" improved significantly at 3 months. |
| 12 | “A group-based lifestyle intervention for diabetes prevention in low-and middle-income country: implementation evaluation of the Kerala Diabetes Prevention Program”(Aziz et al., 2018)” | **Design:** *randomized control trial*<br>**Subject :** 1327 respondents<br>**Variable:**<br>**Independent :** *lifestyle intervention*<br>**Dependent:** *Provider-level factor, participant-level factor, community-level factor*<br>**Instrument :** Questionnaire<br>**Analysis :** Statistical description | Most of the peer leaders participated in the two-day training for peer leaders utilising a manual. Eight sessions were generally attended by the participants. The majority of participants regarded the group sessions to be highly beneficial for changing their lifestyles and were very engaged in them. Inconvenient time and location were identified as the main reasons why participants did not attend all of the sessions. Walking clubs, training for kitchen gardens, and yoga classes were community-based initiatives that were implemented successfully. Peer leaders offer after-class support, and participants report having interaction with their peer leaders outside of class. |
| 13 | “Using Peer Support to Prevent Diabetes: Results of a Pragmatic RCT” (Heisler et al., 2023) | **Design:** *randomized controlled pragmatic clinical trial*<br>**Subject :** 355 respondents<br>**Variable:**<br>**Independent :** *peer support*<br>**Dependent:** : body weight and HbA1c, enrollment in formal diabetes prevention programs, diet, physical activity, social support, *self-efficacy*, motivation,<br>**Instrument :** Questionnaire<br>**Analysis :** , binary logistic regression, t-test | In terms of HbA1c or changes in body weight, there were no differences between the intervention and control groups. Peer groups have been shown to aid in the treatment and prevention of prediabetes. |
| 14 | “The Impact of Physical Activity on the Prevention of Type 2 Diabetes: Evidence and Lessons Learned From the Diabetes Prevention Program, a Long-Standing Clinical Trial Incorporating Subjective and Objective Activity Measures” (Kriska et al., 2021) | **Desain:** *randomized controlled clinical trial*<br>**Subject :** 1783 respondents<br>**Variable:**<br>**Independent :** physical activity<br>**Dependent :** : prevention of Diabetes mellitus type 2<br>**Instrument :** Questionnaire<br>**Analysis :** Kruskal-Wallis and Wilcoxon signed rank tests, Spearman rank | Per 6 MET-hours/week, the incidence of diabetes decreased by 6% after controlling for variables like age, gender, baseline physical activity, and body weight. When compared to metformin or a placebo, the accumulation of physical activity was higher with lifestyle modifications |
| 15 | “Long-term Outcomes of Lifestyle Intervention to Prevent Diabetes in American Indian and Alaska Native Communities: The Special Diabetes Program for Indians Diabetes Prevention Program” (Jiang et al., 2018a)” | **Design:** Cohort study<br>**Subject :** 8652 respondents<br>**Variable:**<br>**Independent :** Lifestyle interventions<br>**Dependent :** prevention of diabetes mellitus<br>**Instrument :** Questionnaire<br>**Analysis :** Paired t test, ANOVA and regression | After the intervention, respondents who had their weight measured saw an average weight loss of 3.8%. Over a six-year period, participants who lost over 5% of their body weight had a decreased risk of acquiring diabetes. |
| 16 | “Short-term effects of lifestyle intervention in the reversion to normoglycemia in people with prediabetes” (Liu et al., 2022). | **Design:** quantitative<br>**Subject :** 2005 respondents<br>**Variable:**<br>**Independent : Lifestyle interventions**<br>**Dependent : normoglycemia**<br>**Instrument :** Questionnaire<br>**Analysis :** chi-square, T-test, logistic regression | Waist circumference decreased in the intervention group, and fasting plasma glucose levels were noticeably higher than in the control group |
| 17 | “Diabetes prevention through digital therapy for highrisk individuals: Study protocol for the Malaysia Diabetes Prevention Programme (MyDiPP)” (Fauzi et al., 2023) | **Design:** *a randomized controlled trial*<br>**Subject :** 100 respondents<br>**Variable:**<br>**Independent : Lifestyle interventions**<br>**Dependent :** prevention of diabetes mellitus<br>**Instrument :** Questionnaire<br>**Analysis : Independent t-test, ANOVA** | Changes in food intake, physical activity levels, and quality of life, as well as changes in body weight and HbA1c levels |
| 18 | “Integrating Diabetes Prevention Education Among Teenagers Involved in Summer Employment: Encouraging Environments for Health in Adolescence (ENHANCE)” (Yazel-Smith et al., 2020) “ | **Design:** *a randomized controlled trial*<br>**Subject :** 168 respondents<br>**Variable :**<br>**Independent : Diabetes prevention education**<br>**Dependent : physical activity, diet, knowledge, self efficacy, depression.**<br>**Instrument : Questionnaire**<br>**Analisis : paired sample t-tests.** | Physical activity and knowledge at the end of the intervention had increased significantly. |

## DISCUSSION

### The Diabetes Prevention Program

The Diabetes Prevention Programme (DPP) aids those who are at high risk of developing diabetes mellitus or who have prediabetes (American Diabetes Association, 2021). One of the key goals of the DPP is to employ lifestyle interventions to stop high-risk people from developing prediabetes or diabetes mellitus (Jiang et al., 2018). Lifestyle interventions can lower the chance of developing diabetes mellitus, according to several randomised controlled trials, including diabetes mellitus prevention programmes (Gilbertson et al., 2019; Kramer et al., 2018; Sampson et al., 2020; Vargas-Ortiz et al., 2020). Weight loss of at least 7% and 150 minutes per week of moderate physical activity are the targets of lifestyle interventions (American Diabetes Association, 2021; Ford et al., 2019)

### Implementation of Diabetes Prevention Program in High-Risk or Prediabetic Individuals

The method used in the Diabetes prevention program is lifestyle intervention through dietary and physical activity (Ford et al., 2019; Kriska et al., 2021). Health education and behavior change strategies are also included in the lifestyle intervention in the Diabetes mellitus prevention program (Soltero et al., 2018). The Diabetes mellitus prevention program through lifestyle intervention is a goal-based intervention where all participants are given the same intervention to achieve the goal of losing weight and doing physical activity, but are allowed to use specific methods to achieve the goal (American Diabetes Association, 2021). The following is an overview of the implementation of the Diabetes mellitus prevention program based on a literature review:

### Dietary

Dietary interventions in diabetes mellitus prevention programmes include reducing total food fat and calories and increasing vegetable and fruit intake (Ford et al., 2019) to prevent type 2 diabetes mellitus for individuals at high risk with a body mass index showing excess body weight or obesity (American Diabetes Association, 2021). One of the goals of diabetes mellitus prevention programme is to reduce body weight by 7% in individuals who are overweight or obese (Kramer et al., 2018). Before implementing nutritional or diet interventions, it is necessary to assess food intake for 3 days to determine the total calorie intake and macronutrient composition, including carbohydrates, fats, and proteins, of a daily intake (Gilbertson et al., 2019).

Face-to-face implementation of dietary or nutritional treatments can be done over a period of 12 months and 22 group sessions (16 core sessions in the first 6 months, followed by 6 maintenance sessions) (Kramer et al., 2018). In addition, it can be carried out online through a 12-month intensive intervention with weekly online modules via a web platform (Lee, Pearl G; Damschroder, Laura J; Holleman, Robert; Moin, Tannaz; Richardson, 2018). It can also be carried out via text messages, which can include information on healthy eating and nutrition (Vargas et al., 2023). Online programmes to prevent diabetes mellitus attracted more participants. On the other hand, online or in-person programmes for preventing diabetes mellitus had a 5% effect on weight loss (Lee, Pearl G.; Damschroder, Laura J.; Holleman, Robert; Moin, Tannaz; Richardson, 2018). Digital therapy is another method that can be used to deliver diabetes mellitus prevention programmes. Participants can access digital applications that include health education, health training, peer groups, and tools for tracking nutritional status, intake, physical activity, body weight, and HbA1c (Fauzi et al., 2023). Additionally, putting into practise diabetes mellitus preventive programmes can be done in clinical and communal contexts (Defosset et al., 2021).

To implement dietary or nutritional interventions and increase physical activity, it is necessary to have the support of the family or community. The assistance offered can take the form of evaluation, knowledge, practical assistance, and emotional support (Soltero et al., 2018). With people who are at high risk for diabetes or prediabetes, nutritional and food counselling is also given to family members. The topics are divided into 6 sections, each lasting around an hour. Processing meal planning, creating healthy plates, and analysing and contrasting food labels are all nutrition-related subjects. According to research (Vargas-Ortiz et al., 2020), family members who participate in diabetes mellitus preventive programmes effectively change their glucose and insulin sensitivity. A type of assistance offered by other patients who have adopted healthy lifestyle modifications and received training in preventing diabetes mellitus is known as peer support. In order to improve behaviour and prevent diabetes mellitus for six months, persons with prediabetes or high risk conditions can receive weekly telephone help from peers (Heisler et al., 2023). Patient-centered counselling strategies can be applied to nutritional or dietary therapies, as well as physical exercise, in the prevention of diabetes mellitus. stimulate decision-making regarding behaviour change, boost motivation for change, engage social support, support problem-solving and help with developing personally tailored goals, action plans, and self-monitoring. Participants established a behaviour modification goal of 7% weight loss if their BMI was higher than 30 (Sampson et al., 2020).

### Physical activity

People with prediabetes benefit from 150 minutes per week of moderate-intensity physical exercise, such as brisk walking (American Diabetes Association, 2021; Kramer et al., 2018; Sampson et al., 2020). A person’s risk of developing diabetes mellitus can be decreased if they have a body mass index (BMI) of greater than 30, 150 minutes per week of moderate-intensity exercise, 2 to 3 sessions per week of muscle-strengthening exercise, and a lower intake of total and saturated fats (Sampson et al., 2020). Physical activity’s primary objective is to lower body weight by about 7% (Kriska et al., 2021). Only 20% of people who engage in physical exercise with energy expenditure comparable to 20 minutes of brisk walking will develop Type 2 diabetes (Arsenault & Despr, 2023). Running sprint interval training (INT), which is performed four times for 30 seconds, can also be used as a form of exercise, as can moderate-intensity continuous training (MICT), which entails walking on a treadmill for 30 minutes while maintaining a heart rate reserve of 45– 55%. Every four weeks, the training session is extended by 10 minutes, resulting in a 60-minute session for the MICT group members. According to Gilbertson et al. (2019), these two physical activities are successful at lowering body weight and body fat mass.

Age and socio-demographic characteristics are among those associated with participation and success in diabetes mellitus prevention programmes, according to a literature review (Devaraj et al., 2021; Venkataramani et al., 2019). According to Venkataramani et al. (2019), more than 25% of individuals have never taken part in a diabetic mellitus preventive programme and have no interest in doing

### Outcomes from the Diabetes Mellitus Prevention Programme

The Diabetes Mellitus Prevention Programme, which is a lifestyle intervention, has both short- and long-term effects on avoiding diabetes mellitus in those with prediabetes or high risk of developing the disease (Jiang et al., 2018a; Liu et al., 2022). After three months of the Diabetes mellitus prevention programme with lifestyle intervention, short-term effects were assessed, and long-term effects were assessed after six to twelve months (Jiang et al., 2018b; Liu et al., 2022). Short-term effects include reduced waist circumference and fasting plasma glucose levels, which were significantly higher in the intervention group compared to the control group (odds ratio 1.32, P = 0.02) (Gilbertson et al., 2019; Lee, Pearl G.; Damschroder, Laura J.; Holleman, Robert; Moin, Tannaz; Richardson, 2018). Increased awareness of food, exercise, and Diabetes Mellitus prevention are some other short-term effects (Yazel-Smith et al., 2020).

The Diabetes mellitus prevention programme with lifestyle intervention had the following long-term effects on participants: >5% weight reduction, 17% weight loss between 3 and 5%, and 47% weight loss of less than 3% (Lee, Pearl G; Damschroder, Laura J; Holleman, Robert; Moin, Tannaz; Richardson, 2018). Participants who lost more than 5% of their body weight had a 64% decreased chance of getting diabetes over the course of six years compared to those who lost between 3-5%. (Jiang et al., 2018) have diabetes. Changes in lipid profiles, insulin sensitivity, enhanced pancreatic cell activity, decreased body fat mass, and HbA1C are additional long-term impacts (Vargas-Ortiz et al., 2020; Kramer et al., 2018). A decrease in total energy intake and an increase in fruit and vegetable consumption were also seen (Ford et al., 2019).

## CONCLUSION

To stop the onset of diabetes mellitus in prediabetic people, lifestyle interventions for diabetes prevention must be implemented. Diet, physical activity, health education, and behaviour modification techniques are all examples of lifestyle interventions. The diet’s major goals are to lower overall fat and food calories while increasing intake of fruits and vegetables. Diabetes mellitus can be halted or delayed with 150 minutes of moderate-intensity exercise five days a week or more, two to three sessions of muscle-strengthening exercise, and a reduction in total and saturated fat intake.

## Data Availability

All data produced in the present work are contained in the manuscript

## ACKNOWLEDGMENTS

Thank you to the nursing lecturers at Airlangga University and friends who have helped write this article.

